# Basal but not complex motor control relies on interhemispheric structural connectivity after stroke

**DOI:** 10.1101/2022.10.04.22280666

**Authors:** Theresa Paul, Valerie M. Wiemer, Lukas Hensel, Matthew Cieslak, Caroline Tscherpel, Christian Grefkes, Scott T. Grafton, Gereon R. Fink, Lukas J. Volz

## Abstract

**Objective:** While ample evidence highlights that the ipsilesional corticospinal tract (CST) plays a crucial role in motor recovery after stroke, very few studies have assessed cortico-cortical motor connections with inconclusive results. Given their unique potential to serve as structural reserve enabling motor network reorganization, the question arises whether cortico-cortical connections may facilitate motor control depending on CST damage.

**Methods:** Diffusion spectrum imaging (DSI) and a novel compartmentwise analysis approach were used to quantify structural connectivity between bilateral cortical core motor regions in chronic stroke patients. Basal and complex motor control were differentially assessed.

**Results:** Both basal and complex motor performance were correlated with structural connectivity between bilateral premotor areas and ipsilesional primary motor cortex (M1) as well as interhemispheric M1-M1 connectivity. While complex motor skills depended on CST integrity, a strong association between M1-M1 connectivity and basal motor control was observed independent of CST integrity especially in patients who underwent substantial motor recovery. Harnessing the informational wealth of cortico-cortical connectivity facilitated the explanation of both basal and complex motor control.

**Interpretation:** We demonstrate for the first time that distinct aspects of cortical structural reserve enable basal and complex motor control after stroke. In particular, recovery of basal motor control is supported via an alternative route through contralesional M1 and non-crossing fibers of the contralesional CST. Our findings help to explain previous conflicting interpretations regarding a vicarious or maladaptive role of the contralesional M1 and highlight the potential of structural connectivity of the cortical motor network as a biomarker post-stroke.

## Introduction

Diffusion MRI (dMRI) is commonly used to characterize white matter (WM) alterations associated with motor impairment following stroke.^1^ It is well-established that the capacity for motor control depends on the “microstructural” integrity of descending motor tracts such as the ipsilesional CST.^2^ At the same time, very little attention has been devoted to cortico-cortical structural connectivity, even though functional imaging studies suggest a pivotal role of interactions between cortical motor areas for motor performance in healthy individuals and stroke patients.^3,4^ Both resting-state functional connectivity and task-based effective connectivity have repeatedly been shown to relate to motor impairment in the acute and chronic stages post-stroke.^5–8^ Given the assumed structure-function relationships,^9^ structural connectivity of this cortical motor network might play a seminal role in motor control after stroke. From a mechanistic perspective, structural cortical connectivity may form the basis for altered network dynamics and hence reflect a patients’ structural reserve enabling motor recovery via functional reorganization.^10^

However, studies on cortico-cortical structure-function relationships remain surprisingly scarce. Existing evidence suggests that motor performance relates to structural connectivity between bilateral primary motor cortex (M1).^11–16^ Studies investigating ipsilesional premotor-M1 connectivity have reported inconclusive findings.^11,17,18^ Moreover, data on the role of interhemispheric premotor-M1 connections are missing. While whole-brain analyses principally include these connections, typical atlas parcellations do not isolate known premotor areas, limiting their interpretability.^19,20^ Moreover, most studies commonly focus on either CST integrity or cortical connectivity. Hence, it remains unknown how post-stroke motor control is facilitated via reorganization based on structural connectivity of the cortical motor network or whether such reorganization depends on the extent of ipsilesional CST damage.

To address these issues, we assessed diffusion spectrum imaging (DSI) data in a sample of chronic stroke patients. Using a novel approach,^21^ compartmentwise generalized fractional anisotropy (gFA) was extracted from specific cortico-cortical tracts defined based on normative HCP data.^22^ Tracts were defined for a network of cortical motor areas. We systematically assessed the relationship of various cortico-cortical connections with basal and complex motor functions in chronic stroke patients. In line with functional and effective connectivity findings, we expected bilateral premotor - ipsilesional M1 and interhemispheric M1-M1 connectivity to be indicative of both basal and complex motor skills. Given that basal motor commands such as lifting the arm against gravity might potentially be compensated via alternative routes such as non-crossing fibers of the contralesional CST,^23^ we hypothesized a stronger dependence on ipsilesional CST integrity for complex than for basal motor skills. Importantly, all analyses were repeated while controlling for ipsilesional CST integrity. Finally, we addressed whether structural connectivity differed in patients with substantial motor recovery compared to patients with limited or no recovery from the acute to the chronic phase post-stroke. This approach allowed us to identify features of cortico-cortical structural connectivity associated with successful motor recovery. Advancing our mechanistic understanding of motor recovery will help to lay the foundation for targeted therapeutic interventions and to identify cortico-cortical connections as potential biomarkers.

## Material and methods

### Subjects

Twenty-five chronic stroke patients (mean age=66.68, std=11.25, 5 female, 20 male) formerly hospitalized at the University Hospital Cologne, Department of Neurology, were included (for detailed demographic and clinical information, see Supplementary Table 1). Inclusion criteria were (1) age between 40 and 90 years, (2) first-ever ischemic stroke more than six months ago and (3) initial unilateral impairment of upper limb motor function. Exclusion criteria were (1) any contraindications to MRI, (2) bihemispheric infarctions, (3) cerebral hemorrhage, (4) reinfarction or other neurological diseases, and (5) persistence of severe aphasia or neglect. All subjects provided informed consent. The study was approved by the ethics committee of the Medical Faculty of the University of Cologne and was conducted in accordance with the declaration of Helsinki. While data from the current patient cohort was included in a previous publication focusing on descending corticospinal and extrapyramidal pathways,^21^ there is no overlap with the current analyses assessing cortico-cortical connectivity.

### Behavioral motor tests

To differentially quantify the impairment of basal and complex motor control involving proximal and distal arm movements, motor impairment was assessed using the Action Research Arm Test (ARAT)^24^ and the Motricity Index (MI)-arm score.^25^ The ARAT probes the execution of activities of daily living and therefore requires the complex interplay of motor synergies, emphasizing distal control of hand motor functions. In contrast, the MI-arm reflects more basal motor control with a focus on proximal and some distal upper limb movements (Fig. 1). The universally used National Institutes of Health Stroke Scale (NIHSS)-arm subscore was used to quantify a patient’s degree of motor recovery from the acute to the chronic stage post-stroke (holding arms 90° against gravity; levels 0: no drift, 1: drift, 2: arm falls before 10 s, 3: no effort against gravity, 4: no movement). *Substantial* recovery was defined as NIHSS-arm improvements of one point or more from the acute to the chronic stage (15 patients). Of note, the absence of a change in the NIHSS-arm score should not be equated with no recovery as the NIHSS cannot capture nuanced differences. In other words, a patient who is able to perform all NIHSS items flawlessly might still have difficulties performing certain aspects of the ARAT. Therefore, patients without an increase in the NIHSS-arm score are summarized as *non-substantial* recovery group (10 patients).

**Figure 1:**
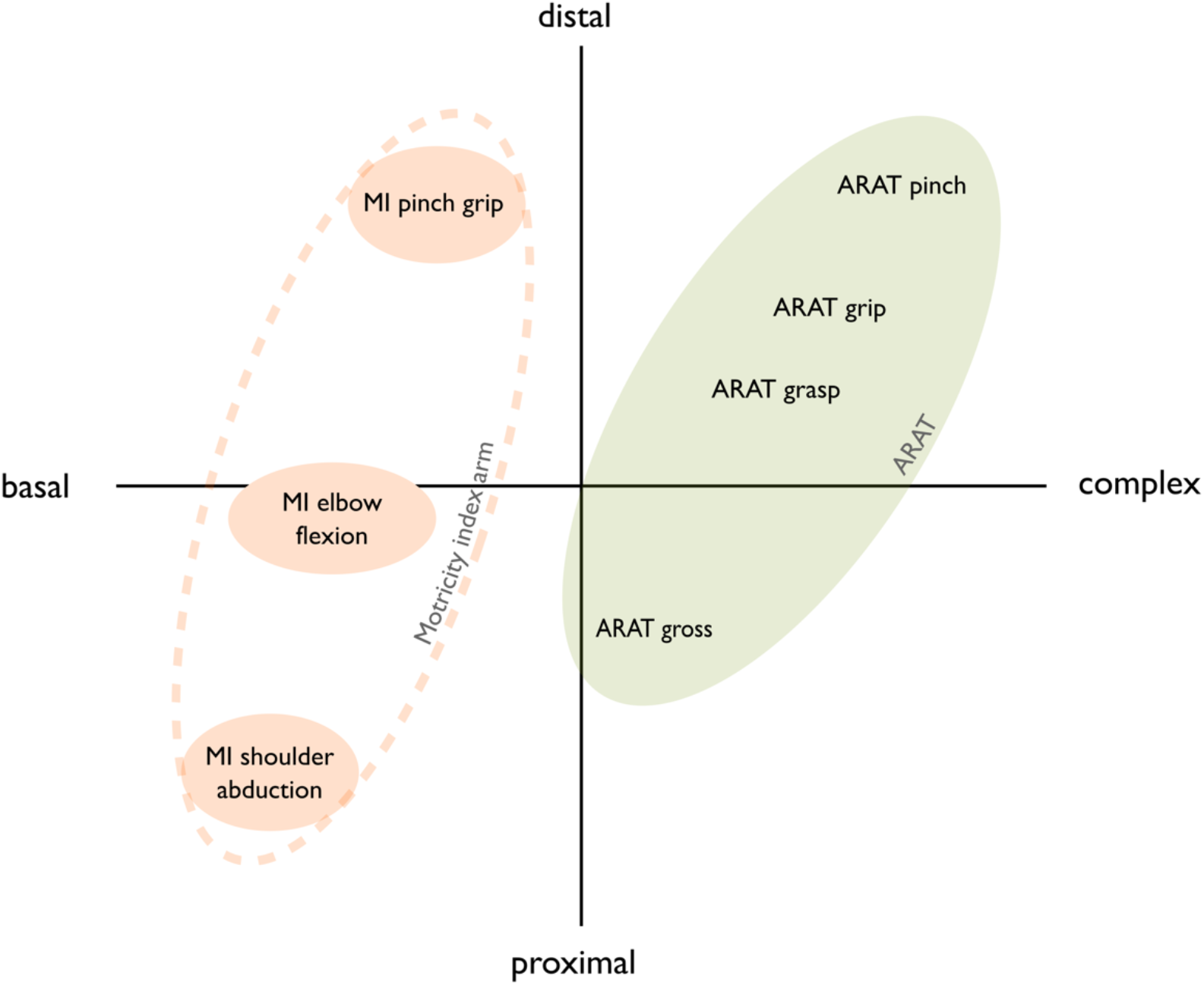
Schematic illustration of motor assessments via the Action Research Arm Test (ARAT) and Motricity Index (MI)-arm score. The MI-arm score (orange) reflects basal motor control of simple movements involving specific muscle synergies with a precise delineation of the reliance on required muscle groups for proximal to distal movements. Conversely, the ARAT (green) quantifies more complex motor control of the affected arm that requires the interplay of different motor control policies, closely reflecting activities of daily living.

### MRI acquisition and preprocessing

MRI data were recorded using a Siemens MAGNETOM Prisma 3 Tesla scanner (Siemens Medical Solutions, Erlangen, Germany). Preprocessing of diffusion data was performed using QSIPrep^26^ and gFA-maps were generated in DSI Studio (https://dsi-studio.labsolver.org/; for a detailed description see^21^). Individual gFA-maps were normalized to MNI-space using ANTS.^27^ Lesion masks were drawn in MRIcron (www.sph.sc.edu/comd/rorden/MRicron) and verified by a certified neurologist. Images with lesions affecting the right hemisphere were flipped along the mid-sagittal plane to facilitate group comparisons. To focus all subsequent analyses on WM voxels and exclude voxels located within the stroke lesion, gFA-maps were masked using both individual WM-masks derived from brain tissue segmentation and lesion masks.

### Defining regions of the cortical motor network

As effective and functional connectivity within a motor network comprising core motor areas have frequently been linked to motor impairment after stroke,^5–7,28^ we accordingly included bilateral core motor areas such as M1, dorsal premotor cortex (PMd), ventral premotor cortex (PMv) and supplementary motor area (SMA). To define the location of the aforementioned areas, term-based fMRI meta-analyses were performed using the neurosynth.org database (https://www.neurosynth.org/) with search terms including “motor cortex”, “dorsal premotor”, “ventral premotor”, and “supplementary motor”. Derived activation patterns were used to define regions of interest (ROIs) for M1 (MNI coordinates left: −38/-22/60, right: 38/-22/60), PMd (left: −24/-6/62, right: 24/-6/62), PMv (left: −54/-1/22, right: 54/-1/22) and SMA (left: −4/-4/54, right: 4/-4/54).

### Generation of tract templates

Fiber bundles connecting cortical motor regions were defined via deterministic fiber tracking as implemented in DSI Studio^29^ using the HCP-1065 template based on diffusion data of 1065 healthy subjects.^22^ Deterministic fiber tracking was used to identify (1) intrahemispheric cortico-cortical fiber tracts between ipsilesional (il) M1 and ipsilesional premotor areas (ilPMd-ilM1, ilPMv-ilM1, ilSMA-ilM1), (2) interhemispheric cortico-cortical fiber tracts between ipsilesional M1 and contralesional (cl) premotor areas (clPMd-ilM1, clPMv-ilM1, clSMA-ilM1), as well as (3) the interhemispheric tract between bilateral M1 (clM1-ilM1; Fig. 2). Fiber tracking was performed using the generated cortical ROIs, exclusion ROIs and an angular threshold of 50-90 degrees. Resulting tracts were manually trimmed and validated by a certified neurologist. To address whether potential associations between structural motor network connectivity and motor impairment were independent of CST integrity, we generated an additional CST mask originating from M1, PMd, PMv and SMA. Importantly, the CST is known to be slightly asymmetrical for the left and right hemispheres in healthy subjects.^30^ Considering that right-hemispheric lesions were flipped to the left hemisphere, left- and right-hemispheric CST tracts were created and combined into a single mask after flipping the right-hemispheric tract along the mid-sagittal plane. Thereby we ensured that all relevant voxels were captured (Fig. 2).

**Figure 2:**
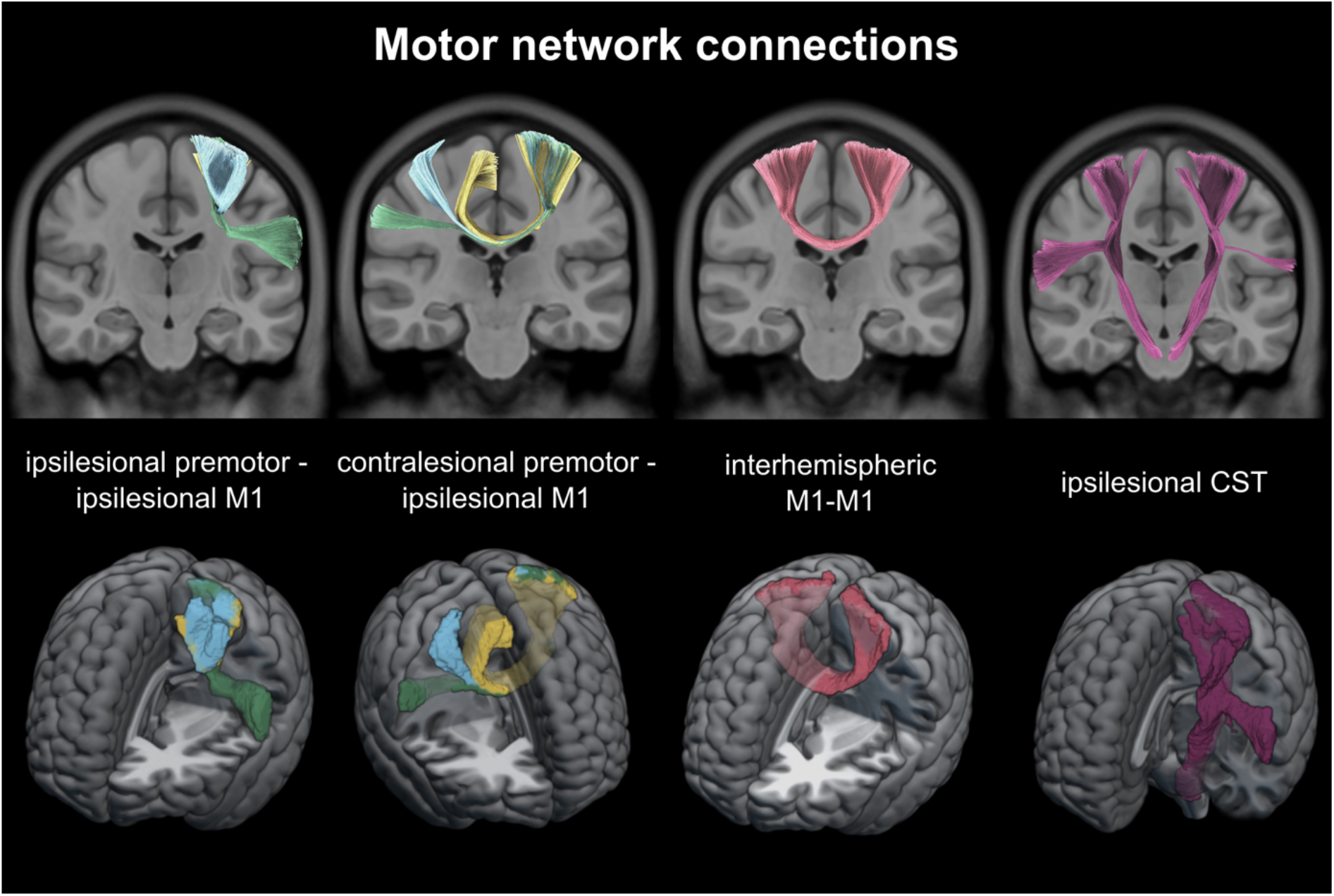
Cortico-cortical and descending motor network connections. Fiber tracts between core areas of the cortical motor network were created using deterministic fiber tracking based on the HCP1065-template22 in DSI Studio (*upper row*). Motor tract templates used for anisotropy extraction are depicted as overlays in MRIcroGL *(lower row)*. Note that for the ipsilesional CST, tracking was first performed in both hemispheres (*upper row*). Bilateral tracts were then combined into a single ipsilesional CST mask after flipping the right-hemispheric tract to the left (*lower row*). *blue = connection with PMd, green = connection with PMv, yellow = connections with SMA, red = interhemispheric M1-M1 connection, purple = corticospinal tract*.

### Tractwise anisotropy

To quantify structural connectivity, diffusion data was compartmentalized using a DSI-based compartmentwise approach.^30^ A deterministic mask was applied to whole-brain gFA-maps in order to differentiate voxels according to the number of trackable fiber directions.^30^ This approach has been shown to facilitate the analyses of anisotropy in stroke patients.^21^ Importantly, tractwise gFA-values were determined based on voxels with only one dominant fiber direction. Focusing the analyses on one-directional voxels helped us to overcome the methodological limitations of biased anisotropy estimations in voxels with multiple fiber directions (for a detailed discussion regarding the impact of compartmentalization on analyses of anisotropy, see ^21,30^).

### Structural connectivity and motor control after stroke

A potential relationship between anisotropy of cortico-cortical motor connections and different aspects of motor control after stroke was tested via Pearson correlations. To probe for relationships with basal motor control, correlations were computed between the MI-arm score and tractwise mean gFA. All p-values were FDR-corrected for multiple comparisons.^31^ To test for relationships with complex motor control, the analyses were repeated using the ARAT score.

To address the question whether the associations between cortico-cortical connectivity and motor control depended on CST damage, partial correlations were computed controlling for ipsilesional CST anisotropy. Importantly, CST integrity has also been related to the degree of motor recovery after stroke.^1^ However, recovery is multifaceted, with good outcomes potentially deriving from various distinct mechanisms at the network level. For example, a small lesion may lead to mild initial impairment which yields a good outcome (almost) independent of the degree of recovery. On the other hand, patients with lesions involving a large amount of brain tissue suffering from severe initial impairment may recover substantially during rehabilitation, also resulting in a good outcome at the chronic stage. Therefore, we assessed the relationship between cortico-cortical motor network connectivity and motor outcome in a recovery-dependent manner. To this end, we divided the patient cohort into two subgroups featuring substantial or non-substantial upper limb recovery, as reflected by improvements in the NIHSS-arm score between the acute and chronic phases. Correlation analyses with basal motor outcome scores were repeated for both subgroups.

To make sure that results were not driven by the direct impact of the lesion, all correlation analyses were repeated after excluding specific tracts in subjects that showed an overlap of more than 10% between the lesion and the tract’s one-directional voxels (N=3).

### Stepwise linear backward regressions

Stepwise linear backward regressions based on the Bayesian information criterion (BIC, k=log(N), N=25) were computed to probe for the extent of explained variance in basal (MI-arm) and complex (ARAT) motor performance by the integrity of ipsilesional CST and cortico-cortical connectivity. For a better appraisal of the ratio of explained variance by the combined model, we separately assessed how much variance was accounted for by (i) cortico-cortical connectivity without the CST and (ii) CST anisotropy alone. As CST damage is considered a valid biomarker for motor impairment post-stroke,^2^ the direct comparison is a good indicator for the suitability of cortico-cortical structural connectivity to potentially improve prediction of behavior.

### Data availability statement

Data are available from the corresponding author upon reasonable request. Tract templates used for the extraction of tractwise anisotropy can be downloaded from the following link: https://tinyurl.com/4ttnnzsr

## Results

### Correlation analyses

For both basal and complex upper limb motor control, positive correlations were observed with anisotropy of the homologous clM1-ilM1 connection, all intrahemispheric premotor-ilM1 connections and interhemispheric clPMv-ilM1 and clSMA-ilM1 connections (all p<.05, FDR-corrected; for details see Table 1 and Fig. 3A). Thus, higher levels of anisotropy were found in patients featuring higher levels of basal and complex motor control of the stroke-affected arm. In general, correlations with tractwise anisotropy tended to be stronger for basal than for complex motor control. Our findings are in line with the notion that structural motor network connectivity between ipsilesional M1 and (i) bilateral premotor areas as well as (ii) contralesional M1 supports both basal and complex motor function of the paretic arm and hand in chronic stroke patients. Considering the prominent role of the CST in motor control, we next addressed the question whether the observed correlations were dependent on the level of CST integrity by means of partial correlations.

**Table 1:**
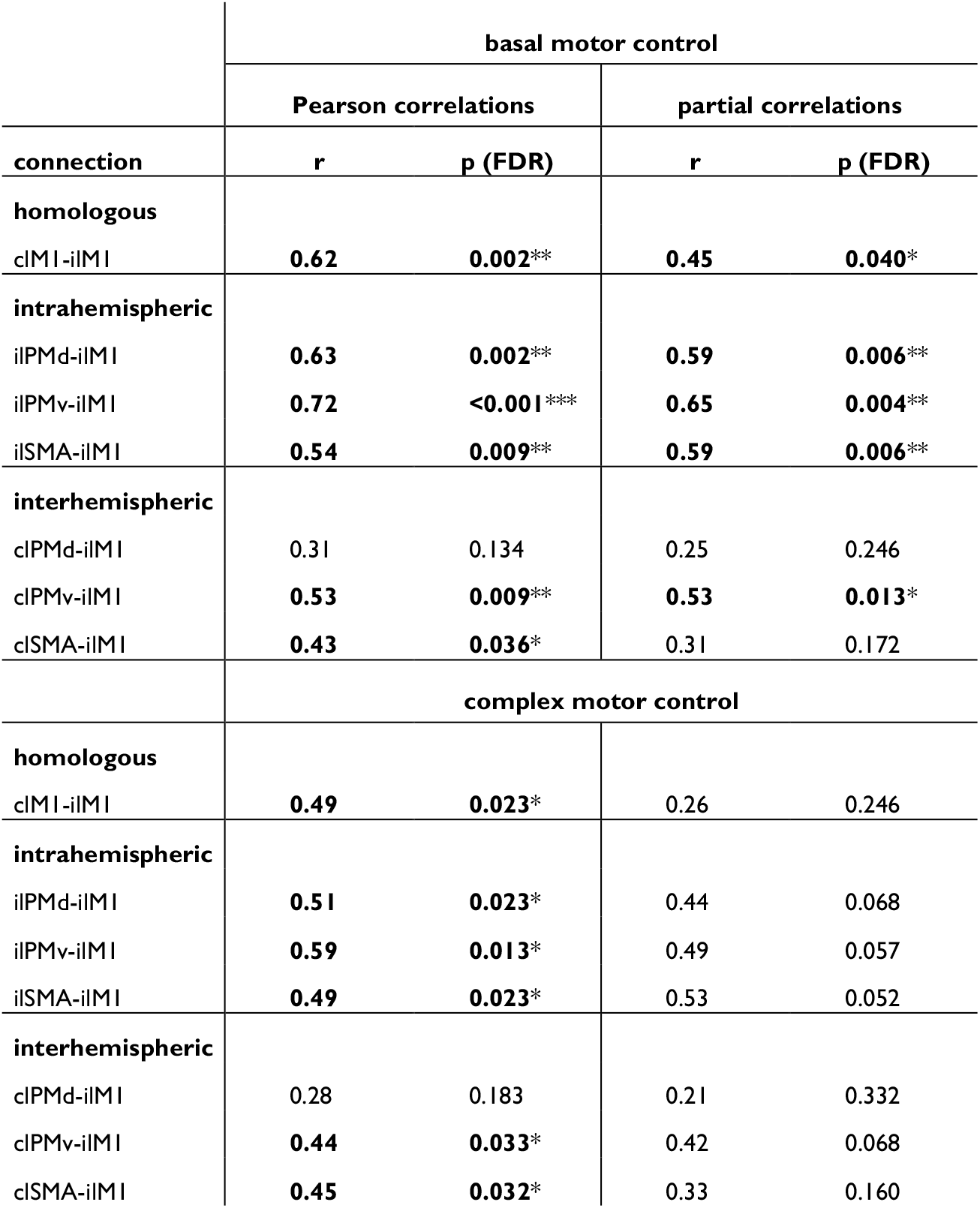
Correlation analyses between different aspects of motor control and cortico-cortical connections. Analyses were carried out separately for (i) basal and (ii) complex motor control. Partial correlations assessed the relationship between motor control and tractwise anisotropy while controlling for ipsilesional CST integrity. Bold font indicates significance after FDR-correction (p < .05). Asterisks signify the following significance thresholds: *** p < .001, ** p < .01, * p < .05. Results are visualized in Fig. 3.

**Figure 3:**
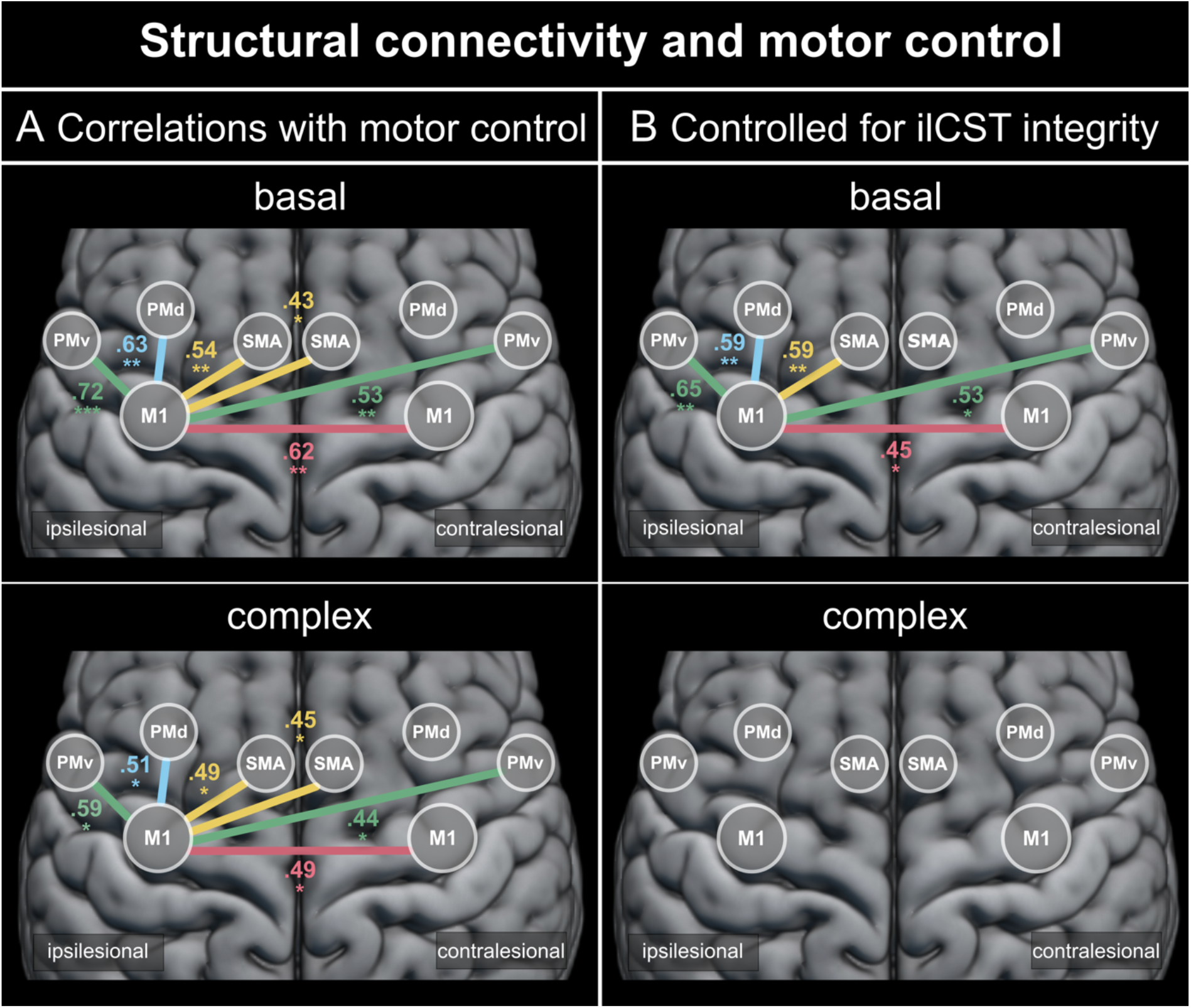
Association between structural motor network connectivity and motor control after stroke. Tractwise anisotropy of several cortico-cortical connections showed a significant association with basal or complex motor control. (**A**) Correlation coefficients of significant Pearson correlations. (**B**) Significant partial correlations of cortico-cortical connections with motor behavior when controlling for ipsilesional CST damage. All depicted connections were significant after FDR-correction for multiple comparisons (p < .05). Significance thresholds: *** p < .001, ** p < .01, * p < .05. *M1 = primary motor cortex, PMd = dorsal premotor cortex, PMv = ventral premotor cortex, SMA = supplementary motor area, ilCST = ipsilesional corticospinal tract*

### Partial correlation analyses

Partial correlation analyses were performed to control for the effect of ipsilesional CST integrity on tractwise correlations with motor behavior. For basal motor control, results of correlation analyses and partial correlations were highly similar (Table 1; Fig. 3). In particular, anisotropy of all intrahemispheric premotor-ilM1 connections (all r>.59, p<.006, FDR-corrected) were associated with basal motor control (Fig. 3B). Regarding interhemispheric connectivity, clPMv-ilM1 (r=.53, p=.013, FDR-corrected) as well as M1-M1 connectivity (r=.45, p=.040, FDR-corrected) also remained significant when controlling for CST integrity. In summary, structural connectivity between the ipsilesional M1 and (i) all ipsilesional premotor areas, (ii) contralesional PMv as well as (iii) contralesional M1 was associated with basal motor control independent of ipsilesional CST integrity.

In contrast, correlations between cortico-cortical connections and complex motor control were not independent of ipsilesional CST integrity (Table 1; Fig. 3B). No significant partial correlations were observed after correction for multiple comparisons. Considering that some premotor-M1 connections showed an FDR-corrected trend towards significance when controlling for CST damage (Table 1), compensation via premotor-M1 connections seemed to show a less pronounced reliance on CST fibers than compensation via the interhemispheric M1-M1 connection. Of note, after excluding lesion-affected tracts, Pearson correlations as well as partial correlations yielded highly similar results, corroborating the robustness of our findings (Supplementary Table 2).

In summary, our findings outline the crucial role of ipsilesional CST integrity for complex motor control after stroke, as compensatory effects at the cortical level seem to be limited in case of substantial ipsilesional CST damage.

### Motor network connectivity and motor recovery

Patients were divided into subgroups with (N=15) and without (N=10) substantial recovery of arm motor function to assess whether the degree of recovery impacted the association between structural motor connectivity and motor control. For patients featuring substantial recovery, basal motor control was strongly associated with mean anisotropy of the homologous clM1-ilM1 tract (r=.79, p=.003, FDR-corrected; Table 2; Fig. 4A) but showed no significant association with premotor-M1 connectivity. Importantly, this association persisted when controlling for CST integrity (r=.75, p=.016, FDR-corrected).

**Table 2:** Recovery-dependent subgroup analysis: Correlations analyses between basal motor control and cortico-cortical connections. Analyses were carried out separately for patients featuring (i) substantial or (ii) no substantial recovery as assessed by the difference in NIHSS-arm score in the acute and chronic stage. Partial correlations assessed the relationship between basal motor control and tractwise anisotropy while controlling for ipsilesional CST integrity. Bold font indicates significance after FDR-correction (p < .05). Asterisks signify the following significance thresholds: *** p < .001, ** p < .01, * p < .05. Results are visualized in Fig. 4.

| connection | substantial recovery |  |  |  |
| --- | --- | --- | --- | --- |
|  | Pearson correlations |  | partial correlations |  |
|  | r | p (FDR) | r | p (FDR) |
| **homologous** |  |  |  |  |
| cMI-iiMI | **0.79** | **0.003\*\*** | **0.75** | **0.016\*** |
| **intrahemispheric** |  |  |  |  |
| iPMd-iiMI | 0.25 | 0.492 | 0.30 | 0.427 |
| iPMv-iiMI | 0.58 | 0.081 | 0.55 | 0.153 |
| iSMA-iiMI | 0.05 | 0.862 | 0.19 | 0.549 |
| **interhemispheric** |  |  |  |  |
| cPMd-iiMI | 0.22 | 0.492 | 0.18 | 0.549 |
| cPMv-iiMI | 0.31 | 0.467 | 0.34 | 0.416 |
| cSMA-iiMI | 0.50 | 0.139 | 0.42 | 0.325 |
|  | **no substantial recovery** |  |  |  |
| **homologous** |  |  |  |  |
| cMI-iiMI | 0.44 | 0.279 | 0.17 | 0.657 |
| **intrahemispheric** |  |  |  |  |
| iPMd-iiMI | **0.85** | **0.007\*\*** | 0.77 | 0.053 |
| iPMv-iiMI | **0.81** | **0.010\*** | 0.73 | 0.060 |
| iSMA-iiMI | **0.90** | **0.003\*\*** | **0.87** | **0.015\*** |
| **interhemispheric** |  |  |  |  |
| cPMd-iiMI | 0.33 | 0.392 | 0.34 | 0.521 |
| cPMv-iiMI | **0.71** | **0.036\*** | 0.68 | 0.076 |
| cSMA-iiMI | 0.30 | 0.392 | 0.21 | 0.657 |

**Figure 4:**
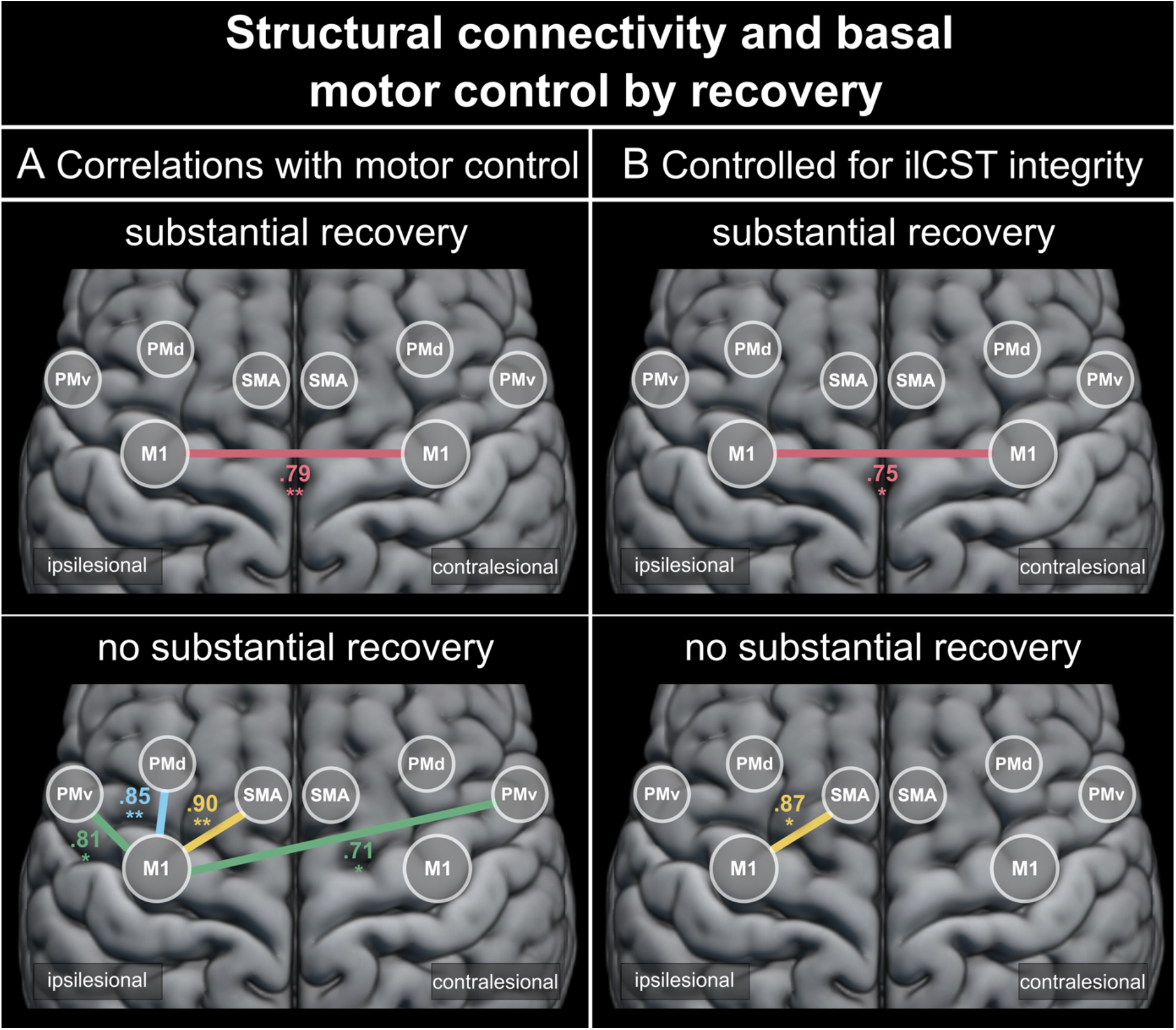
Recovery-dependent subgroup analysis: associations between structural motor network connectivity and basal motor control after stroke. (**A**) Significant Pearson correlations between tractwise anisotropy and basal or complex motor control for patients showing (i) substantial or (ii) no substantial recovery. (**B**) Significant partial correlations between tractwise anisotropy and basal motor control when controlling for ipsilesional CST integrity in patients featuring (i) substantial or (ii) no substantial recovery. Of note, only subjects featuring substantial recovery of arm motor function showed an association of interhemispheric M1-M1 connectivity with basal motor control, highlighting a compensatory role of transcallosal fibers. All depicted connections were significant after FDR-correction for multiple comparisons (p < .05). Significance thresholds: *** p < .001, ** p < .01, * p < .05. *M1 = primary motor cortex, PMd = dorsal premotor cortex, PMv = ventral premotor cortex, SMA = supplementary motor area, ilCST = ipsilesional corticospinal tract*

Conversely, for patients showing no substantial recovery of arm motor function, the interhemispheric clPMv-ilM1 connection (r=.71, p=.036, FDR-corrected) as well as intrahemispheric premotor-ilM1 connectivity was significantly correlated with motor control (all r>.81, p<.011, FDR-corrected; Table 2; Fig. 4A). Of note, no significant association was observed for the clM1-ilM1 connection in patients without substantial recovery (r=.44, p=.279, FDR-corrected). When controlling for CST integrity, only the ilSMA-ilM1 connection yielded a significant correlation (r=.87, p=.015, FDR-corrected). Again, excluding directly affected cortico-cortical connections yielded highly similar results (Supplementary Table 3).

In summary, while patients with substantial motor recovery seemed to heavily rely on interhemispheric M1-M1 connectivity to ensure basal motor control, patients without substantial recovery of arm function featured no such association. Hence, our findings highlight an essential role of interhemispheric M1-M1 connectivity in motor recovery, which may serve as a critical route to recruit the intact contralesional motor network and its descending pathways to compensate for the lesion-induced impairment of motor control after stroke.

### Stepwise linear backward regressions

First, we assessed the propensity of CST integrity to explain behavioral impairment. For basal motor control, 28% of variance (R^2^ = 27.71%, adjusted R^2^= 24.57%, p=.007, BIC=132.46) was explained by the ipsilesional CST. A highly similar result was obtained for complex motor control (R^2^=27.77%, adjusted R^2^=24.63%, p=.007, BIC=130.88). Stepwise backward regression models including cortico-cortical connections and ipsilesional CST integrity explained a high amount of variance for both basal and complex motor control. Specifically, 71% of variance in basal motor control (R^2^=71.01%, adjusted R^2^=65.22%, p<.001, BIC=119.27) and 60% of variance in complex motor control (R^2^=60.17%, adjusted R^2^=46.90%, p=.006, BIC=132.09) were captured. When excluding the ipsilesional CST and only using cortico-cortical connections to start the backward elimination process, the resulting model still explained a substantial amount of variance in basal motor control (R^2^=63.26%, adjusted R^2^=58.01%, p<.001, BIC=121.98). Conversely, while complex motor control was still explained significantly (R^2^=35.02%, adjusted R^2^=32.19%, p=.002, BIC=128.24), the ratio of explained variance in complex motor control considerably decreased after ipsilesional CST exclusion. Thus, the stepwise linear backward regression analyses showed that structural motor network connectivity holds valuable information on motor control across subjects. While basal motor control is readily explained by cortico-cortical connectivity, complex motor control crucially relies on ipsilesional CST integrity.

## Discussion

Motor control is assumed to rely on a distributed network of cortical and subcortical motor areas, as well as its descending pathways such as the CST.^32^ At the cortical level, premotor areas are crucially involved in shaping motor commands in M1 via dense cortico-cortical connections.^33^ After stroke-inflicted CST damage, the question arises how the motor network can reorganize its functional architecture to recover motor control. From a mechanistic perspective, stronger structural cortico-cortical connections may allow for a more flexible and efficient transmission of motor signals, thereby enabling increased influences of bilateral premotor areas onto ipsilesional M1.^5^ Alternatively, retrograde coupling from M1 onto premotor areas might also play a role in compensating motor control. In this case, output signals that cannot be transmitted through damaged CST fibers originating from M1 might be relayed via CST fibers descending from premotor areas or non-crossing contralesional CST fibers originating from contralesional M1. Of note, the interplay and functional significance of these proposed mechanisms remains largely unknown and has resulted in conflicting interpretations of previous findings. Given that recovery of basal and complex movements has been shown to differentially benefit from rehabilitation,^34^ we hypothesized that both derive from distinct mechanisms.

### Complex motor control

After stroke, intact structural connections are thought to enable the functional reorganization of motor network dynamics to facilitate recovery. In line with this assumption, we here observed significant correlations between complex motor skills and cortico-cortical connectivity from contralesional M1 and bilateral premotor areas to the ipsilesional M1 (Fig. 3A). However, when controlling for ipsilesional CST integrity, no significant correlations were observed between cortico-cortical motor connections and complex motor control (Fig. 3B). In other words, while partial correlations with intrahemispheric premotor-M1 connectivity showed a trend towards significance (Table 1), similar variance in complex motor control was captured by anisotropy of ipsilesional CST and cortico-cortical connectivity. An explanation for this observation may derive from previous results indicating that motor performance post-stroke critically relies on the task-dependent modulation of ipsilesional premotor-M1 connectivity.^5,7^ Hence, premotor areas may use intact structural cortico-cortical connections to enhance recruitment of the ipsilesional M1. However, the potential to adapt hand motor output signals on the cortical level may be critically limited in case of extensive CST damage, stressing the significance of ipsilesional CST integrity. This notion is well in line with earlier studies frequently reporting correlations between ipsilesional CST integrity and hand motor function.^2,35–38^ Moreover, our stepwise regression analyses performed best when including CST anisotropy in the starting model. In other words, the CST explained relevant behavioral variance in addition to the information contained in cortico-cortical connectivity. Thus, complex motor commands seem to critically rely on ipsilesional CST output signals.

### Basal motor control

We conceptualized basal motor control as simple movements that require only basic control of specific muscle synergies such as lifting the arm against gravity. While previous diffusion imaging studies have not typically differentiated basal and complex motor control, more basal motor functions have previously been associated with varying features of ipsilesional premotor-M1^11,17,18^ as well as transcallosal M1-M1^11–14^ structural connectivity, painting a rather inconclusive picture. We here observed associations of the MI-arm with anisotropy of all three ipsilesional premotor-M1 connections, interhemispheric premotor-M1 connectivity, and interhemispheric M1-M1 connectivity (Fig. 3A). From a mechanistic perspective, stronger structural cortico-cortical connections may allow for a more flexible and efficient transmission of motor signals, thereby enabling increased influences of bilateral premotor areas onto ipsilesional M1.^5^ Alternatively, retrograde coupling from M1 onto premotor areas might also play a role in compensating basal motor skills. In this case, output signals that cannot be transmitted through damaged CST fibers originating from M1 might be relayed via CST fibers descending from premotor areas. Of note, partial correlation analyses keeping the influence of ipsilesional CST anisotropy constant revealed that associations between cortico-cortical connections and basal motor control were largely independent of CST integrity (Fig. 3B). In other words, basal motor performance showed less reliance on CST anisotropy than complex motor control. The fact that anisotropy between bilateral M1 was significantly associated with basal motor control but not complex motor control indicates that basal but not complex motor skills may be compensated via recruitment of the contralesional M1. Importantly, the contralesional M1 has access to alternative descending routes such as the intact contralesional CST which may relay motor output signals from ipsilesional M1.^23^

Our findings thus highlight a differential role of interhemispheric M1-M1 structural connectivity for basal and complex motor control which helps to shed light on the heavily debated role of the contralesional M1. While some authors have argued that the contralesional M1 serves a vicarious role by contributing to motor control of the paretic hand,^39–42^ other results favor a maladaptive influence of the contralesional M1.^43–45^ Maladaptation is frequently conceptualized to result from increased inhibitory influences exerted by the contralesional M1 onto the ipsilesional M1 as demonstrated by transcranial magnetic stimulation^44^ and fMRI-based effective connectivity.^43,46^

Conversely, the contralesional M1 may play a supportive role by (i) exerting facilitatory influences on the ipsilesional M1^5^ or by (ii) offering an alternative route for descending motor commands via the contralesional CST. As non-crossing CST fibers predominantly innervate proximal arm and shoulder muscles,^32^ this pathway is situated to support proximal arm and shoulder movements rather than fine motor control of the hand and fingers. Our current findings are perfectly in line with this notion as basal motor control involving proximal muscle groups but not complex motor control was associated with interhemispheric connectivity.

Moreover, our recovery-dependent results emphasize that interhemispheric M1-M1 connectivity constitutes a structural reserve for the reorganization of basal motor control. Patients featuring substantial recovery of motor function showed strong correlations between interhemispheric M1-M1 connectivity and basal motor control independent of ipsilesional CST integrity (Fig. 4). Support for this notion stems from Stewart and colleagues who reported anisotropy of callosal motor regions to be linked to (basal) motor control in patients with favorable motor outcomes at the chronic stage.^14^ Hence, recovery of basal motor function seems to depend on a patient’s ability to recruit the contralesional M1 via interhemispheric callosal fibers.

### Clinical relevance

Our current findings highlight the potential of structural cortico-cortical motor network connectivity as a biomarker to predict motor impairment following stroke. Structural scans can be easily integrated into the clinical routine as they only require little patient compliance.^47,48^ For example, the DSI scanning protocol used in the present study offers a fast acquisition time of only 11 minutes. However, the reliable assessment of cortical-cortical connectivity via anisotropy is hindered by voxels with multiple fiber directions. To overcome this problem we employed a compartmentwise approach that differentiates voxels according to the number of trackable fiber directions.^30^ Importantly, stepwise backward regression analyses yielded a high ratio of explained variance for both basal and complex motor skills, by far exceeding the ratio of explained variance achieved by CST integrity alone. Adding CST integrity as a variable to the starting model of the stepwise backward regression drastically increased the percentage of explained variance for complex motor control yet only yielded little additional explained variance for basal motor performance. Thus, a potential biomarker should ideally be task-specific and focus on cortico-cortical connectivity for basal motor skills while including cortico-cortical connectivity and CST integrity in concert for complex motor skills.

### Limitations

A major limitation pertains to the limited sample size of 25 chronic stroke patients. While this sample size is not unusual for hypothesis-driven imaging studies in patient cohorts, a larger sample size would allow for additional analyses. Moreover, one might argue that our results may have been biased by lesions affecting cortico-cortical connections. However, most patients featured subcortical lesions that primarily affected the internal capsule. Thus, cortico-cortical tracts were hardly ever directly affected. Moreover, repeating the analyses without subjects showing a significant lesion overlap with specific cortico-cortical connections yielded highly similar results. Thus, a considerable lesion-induced bias seems unlikely.

## Conclusion

Our current findings highlight the seminal importance of structural cortico-cortical motor network connectivity which serves as a structural reserve for distinct aspects of motor control post-stroke. Our data emphasize that complex motor control depends on an interplay of cortico-cortical information integration and descending motor commands via the ipsilesional CST. Thus, severe CST damage seems to preclude the control of complex motor functions of the paretic hand. Conversely, basal motor control can be successfully compensated via alternative routes: Interhemispheric pathways between bilateral M1 seem to play a crucial role in relaying motor commands to the contralesional motor cortex. This might help to access intact descending pathways such as non-crossing fibers of the contralesional CST.^23^ Especially patients who underwent substantial recovery from the acute to the chronic stage post-stroke seemed to heavily rely on this route emphasizing its seminal role in functional reorganization of basal motor control. Finally, our results identified a combination of cortico-cortical structural connectivity and CST integrity as a possible biomarker for basal and complex motor functions after stroke which is emphasized by the high degree of explained behavioral variance when using both in concert.

## Supporting information

STROBE_checklist_Paul_et_al

## List of abbreviations

ARAT: Action Research Arm Test
BIC: Bayesian information criterion
cl: contralesional
CST: corticospinal tract
dMRI: diffusion magnetic resonance imaging
DSI: diffusion spectrum imaging
FDR: false discovery rate
gFA: generalized fractional anisotropy
il: ipsilesional
M1: primary motor cortex
MI: Motricity Index
NIHSS: National Institutes of Health Stroke Scale
PMd: dorsal premotor cortex
PMv: ventral premotor cortex
ROI: region of interest
SMA: supplementary motor area
WM: white matter

## Acknowledgements

GRF gratefully acknowledges support from the Marga and Walter Boll Stiftung, Kerpen, Germany. CG and GRF are funded by the Deutsche Forschungsgemeinschaft (DFG, German Research Foundation) – Project-ID 431549029 – SFB 1451. STG was supported by the Institute for Collaborative Biotechnologies under Cooperative Agreement W911NF-19-2-0026 with the Army Research Office.

## Author contributions

TP, CG, STG, GRF and LJV contributed to the conception and design of the study. TP, VMW, LH and CT made substantial contributions to the acquisition of the data. TP, VMW, MC and LJV contributed to the analysis of the data. TP, VMW, STG and LJV contributed significantly to drafting the manuscript and figures. All authors have critically revised and finally approved the version of the paper to be published.

## Supplementary material

Supplementary material is available online.

## References

1. Koch P, Schulz R, Hummel FC. Structural connectivity analyses in motor recovery research after stroke. Ann Clin Transl Neurol. 2016;3(3):233–244. doi:10.1002/acn3.278

2. Boyd LA, Hayward KS, Ward NS, et al. Biomarkers of stroke recovery: Consensus-based core recommendations from the Stroke Recovery and Rehabilitation Roundtable. Int J Stroke. 2017;12(5):480–493. doi:10.1177/1747493017714176

3. Pool EM, Rehme AK, Fink GR, Eickhoff SB, Grefkes C. Network dynamics engaged in the modulation of motor behavior in healthy subjects. Neuroimage. 2013;82:68–76. doi:10.1016/j.neuroimage.2013.05.123

4. Grefkes C, Ward NS. Cortical Reorganization After Stroke. Neurosci. 2014;20(1):56–70. doi:10.1177/1073858413491147

5. Rehme AK, Eickhoff SB, Wang LE, Fink GR, Grefkes C. Dynamic causal modeling of cortical activity from the acute to the chronic stage after stroke. Neuroimage. 2011;55(3):1147–1158. doi:10.1016/j.neuroimage.2011.01.014

6. Rehme AK, Grefkes C. Cerebral network disorders after stroke: evidence from imaging-based connectivity analyses of active and resting brain states in humans. J Physiol. 2013;591(1):17–31. doi:10.1113/jphysiol.2012.243469

7. Paul T, Hensel L, Rehme AK, et al. Early motor network connectivity after stroke: An interplay of general reorganization and state-specific compensation. Hum Brain Mapp. 2021;42(16):5230–5243. doi:10.1002/hbm.25612

8. Golestani AM, Tymchuk S, Demchuk A, Goodyear BG. Longitudinal Evaluation of Resting-State fMRI After Acute Stroke With Hemiparesis. Neurorehabil Neural Repair. 2013;27(2):153–163. doi:10.1177/1545968312457827

9. Rubinov M, Sporns O. Complex network measures of brain connectivity: Uses and interpretations. Neuroimage. 2010;52(3):1059–1069. doi:10.1016/j.neuroimage.2009.10.003

10. Di Pino G, Pellegrino G, Assenza G, et al. Modulation of brain plasticity in stroke: A novel model for neurorehabilitation. Nat Rev Neurol. 2014;10(10):597–608. doi:10.1038/nrneurol.2014.162

11. Peters DM, Fridriksson J, Stewart JC, et al. Cortical disconnection of the ipsilesional primary motor cortex is associated with gait speed and upper extremity motor impairment in chronic left hemispheric stroke. Hum Brain Mapp. 2018;39(1):120–132. doi:10.1002/hbm.23829

12. Chen JL, Schlaug G. Resting State Interhemispheric Motor Connectivity and White Matter Integrity Correlate with Motor Impairment in Chronic Stroke. Front Neurol. 2013;4. doi:10.3389/fneur.2013.00178

13. Hayward K, Ferris JK, Lohse KR, et al. Observational Study of Neuroimaging Biomarkers of Severe Upper Limb Impairment After Stroke. Neurology. Published online May 12, 2022:1–42. doi:10.1212/WNL.0000000000200517

14. Stewart JC, Dewanjee P, Tran G, et al. Role of corpus callosum integrity in arm function differs based on motor severity after stroke. NeuroImage Clin. 2017;14:641–647. doi:10.1016/j.nicl.2017.02.023

15. Wang LE, Tittgemeyer M, Imperati D, et al. Degeneration of corpus callosum and recovery of motor function after stroke: A multimodal magnetic resonance imaging study. Hum Brain Mapp. 2012;33(12):2941–2956. doi:10.1002/hbm.21417

16. Lindenberg R, Zhu LL, Rüber T, Schlaug G. Predicting functional motor potential in chronic stroke patients using diffusion tensor imaging. Hum Brain Mapp. 2012;33(5):1040–1051. doi:10.1002/hbm.21266

17. Schulz R, Braass H, Liuzzi G, et al. White matter integrity of premotor–motor connections is associated with motor output in chronic stroke patients. NeuroImage Clin. 2015;7:82–86. doi:10.1016/j.nicl.2014.11.006

18. Schulz R, Park E, Lee J, et al. Interactions Between the Corticospinal Tract and Premotor–Motor Pathways for Residual Motor Output After Stroke. Stroke. 2017;48(10):2805–2811. doi:10.1161/STROKEAHA.117.016834

19. Schlemm E, Schulz R, Bönstrup M, et al. Structural brain networks and functional motor outcome after stroke—a prospective cohort study. Brain Commun. 2020;2(1):1–13. doi:10.1093/braincomms/fcaa001

20. Kalinosky BT, Schindler-Ivens S, Schmit BD. White matter structural connectivity is associated with sensorimotor function in stroke survivors. NeuroImage Clin. 2013;2(1):767–781. doi:10.1016/j.nicl.2013.05.009

21. Paul T, Cieslak M, Hensel L, et al. The role of corticospinal and extrapyramidal pathways in motor impairment after stroke. Brain Commun. Published online 2022. doi:10.1093/braincomms/fcac301

22. Yeh FC, Panesar S, Fernandes D, et al. Population-averaged atlas of the macroscale human structural connectome and its network topology. Neuroimage. 2018;178(April):57–68. doi:10.1016/j.neuroimage.2018.05.027

23. Schaechter JD, Fricker ZP, Perdue KL, et al. Microstructural status of ipsilesional and contralesional corticospinal tract correlates with motor skill in chronic stroke patients. Hum Brain Mapp. 2009;30(11):3461–3474. doi:10.1002/hbm.20770

24. Lyle RC. A performance test for assessment of upper limb function in physical rehabilitation treatment and research. Int J Rehabil Res. 1981;4(4):483–492. doi:10.1097/00004356-198112000-00001

25. Demeurisse G, Demol O, Robaye E. Motor Evaluation in Vascular Hemiplegia. Eur Neurol. 1980;19(6):382–389. doi:10.1159/000115178

26. Cieslak M, Cook PA, He X, et al. QSIPrep: an integrative platform for preprocessing and reconstructing diffusion MRI data. Nat Methods. 2021;18(7):775–778. doi:10.1038/s41592-021-01185-5

27. Avants BB, Epstein CL, Grossman M, Gee JC. Symmetric diffeomorphic image registration with cross-correlation: evaluating automated labeling of elderly and neurodegenerative brain. Med Image Anal. 2008;12(1):26–41. doi:10.1016/j.media.2007.06.004

28. Volz LJ, Sarfeld AS, Diekhoff S, et al. Motor cortex excitability and connectivity in chronic stroke: a multimodal model of functional reorganization. Brain Struct Funct. 2015;220(2):1093–1107. doi:10.1007/s00429-013-0702-8

29. Yeh FC, Verstynen TD, Wang Y, Fernández-Miranda JC, Tseng WYI. Deterministic diffusion fiber tracking improved by quantitative anisotropy. PLoS One. 2013;8(11):1–16. doi:10.1371/journal.pone.0080713

30. Volz LJ, Cieslak M, Grafton ST. A probabilistic atlas of fiber crossings for variability reduction of anisotropy measures. Brain Struct Funct. 2018;223(2):635–651. doi:10.1007/s00429-017-1508-x

31. Benjamini Y, Hochberg Y. Controlling the False Discovery Rate: A Practical and Powerful Approach to Multiple Testing. J R Stat Soc Ser B. 1995;57(1):289–300. doi:10.1111/j.2517-6161.1995.tb02031.x

32. Lemon RN. Descending Pathways in Motor Control. Annu Rev Neurosci. 2008;31(1):195–218. doi:10.1146/annurev.neuro.31.060407.125547

33. Strick PL, Dum RP, Rathelot JA. The Cortical Motor Areas and the Emergence of Motor Skills: A Neuroanatomical Perspective. Annu Rev Neurosci. 2021;44(1):425–447. doi:10.1146/annurev-neuro-070918-050216

34. George SH, Rafiei MH, Borstad A, Adeli H, Gauthier L V. Gross motor ability predicts response to upper extremity rehabilitation in chronic stroke. Behav Brain Res. 2017;333(June):314–322. doi:10.1016/j.bbr.2017.07.002

35. Puig J, Blasco G, Daunis-I-Estadella J, et al. Decreased Corticospinal Tract Fractional Anisotropy Predicts Long-term Motor Outcome After Stroke. Stroke. 2013;44(7):2016–2018. doi:10.1161/STROKEAHA.111.000382

36. Stinear CM, Barber PA, Smale PR, Coxon JP, Fleming MK, Byblow WD. Functional potential in chronic stroke patients depends on corticospinal tract integrity. Brain. 2007;130(Pt 1):170–180. doi:10.1093/brain/awl333

37. Lindenberg R, Renga V, Zhu LL, Betzler F, Alsop D, Schlaug G. Structural integrity of corticospinal motor fibers predicts motor impairment in chronic stroke. Neurology. 2010;74(4):280–287. doi:10.1212/WNL.0b013e3181ccc6d9

38. Peters DM, Fridriksson J, Richardson JD, et al. Upper and Lower Limb Motor Function Correlates with Ipsilesional Corticospinal Tract and Red Nucleus Structural Integrity in Chronic Stroke: A Cross-Sectional, ROI-Based MRI Study. Tambasco N, ed. Behav Neurol. 2021;2021:1–10. doi:10.1155/2021/3010555

39. Biernaskie J, Szymanska A, Windle V, Corbett D. Bi-hemispheric contribution to functional motor recovery of the affected forelimb following focal ischemic brain injury in rats. Eur J Neurosci. 2005;21(4):989–999. doi:10.1111/j.1460-9568.2005.03899.x

40. Johansen-Berg H, Rushworth MFS, Bogdanovic MD, Kischka U, Wimalaratna S, Matthews PM. The role of ipsilateral premotor cortex in hand movement after stroke. Proc Natl Acad Sci. 2002;99(22):14518–14523. doi:10.1073/pnas.222536799

41. Lotze M, Markert J, Sauseng P, Hoppe J, Plewnia C, Gerloff C. The role of multiple contralesional motor areas for complex hand movements after internal capsular lesion. J Neurosci. 2006;26(22):6096–6102. doi:10.1523/JNEUROSCI.4564-05.2006

42. Rehme AK, Fink GR, von Cramon DY, Grefkes C. The Role of the Contralesional Motor Cortex for Motor Recovery in the Early Days after Stroke Assessed with Longitudinal fMRI. Cereb Cortex. 2011;21(4):756–768. doi:10.1093/cercor/bhq140

43. Grefkes C, Nowak DA, Eickhoff SB, et al. Cortical connectivity after subcortical stroke assessed with functional magnetic resonance imaging. Ann Neurol. 2008;63(2):236–246. doi:10.1002/ana.21228

44. Murase N, Duque J, Mazzocchio R, Cohen LG. Influence of interhemispheric interactions on motor function in chronic stroke. Ann Neurol. 2004;55(3):400–409. doi:10.1002/ana.10848

45. Takeuchi N, Izumi SI. Maladaptive Plasticity for Motor Recovery after Stroke: Mechanisms and Approaches. Neural Plast. 2012;2012:1–9. doi:10.1155/2012/359728

46. Grefkes C, Nowak DA, Wang LE, Dafotakis M, Eickhoff SB, Fink GR. Modulating cortical connectivity in stroke patients by rTMS assessed with fMRI and dynamic causal modeling. Neuroimage. 2010;50(1):233–242. doi:10.1016/j.neuroimage.2009.12.029

47. Horn U, Grothe M, Lotze M. MRI Biomarkers for Hand-Motor Outcome Prediction and Therapy Monitoring following Stroke. Neural Plast. 2016;2016:1–12. doi:10.1155/2016/9265621

48. Bonkhoff AK, Grefkes C. Precision medicine in stroke: towards personalized outcome predictions using artificial intelligence. Brain. 2022;145(2):457–475. doi:10.1093/brain/awab439

