## Supplementary_materials for "Basal but not complex motor control relies on interhemispheric structural connectivity after stroke"

### Supplement

**Supplementary Table 1: Demographic and clinical patient information.** *f = female, m = male, l = left, r = right, MCA = middle cerebral artery, PCA = posterior cerebral artery, ACA = anterior cerebral artery*

| patient | sex | lesion side | lesion location | ARAT | MI-arm | NIHSS-arm acute | NIHSS-arm chronic | substantial recovery | months since stroke |
|---|---|---|---|---|---|---|---|---|---|
| 1 | m | r | MCA (subcortical) | 57 | 99 | 2 | 0 | yes | 21 |
| 2 | m | r | MCA (subcortical) | 57 | 91 | 4 | 1 | yes | 14 |
| 3 | m | r | MCA (subcortical) | 38 | 76 | 1 | 1 | no | 17 |
| 4 | m | l | MCA (subcortical) | 0 | 34 | 3 | 4 | no | 51 |
| 5 | m | l | PCA (subcortical) | 35 | 92 | 1 | 1 | no | 23 |
| 6 | f | l | MCA (subcortical) | 32 | 77 | 1 | 2 | no | 11 |
| 7 | m | l | MCA (subcortical) | 19 | 65 | 3 | 2 | yes | 59 |
| 8 | m | l | MCA (subcortical) | 55 | 92 | 4 | 1 | yes | 71 |
| 9 | m | l | ACA/MCA (subcortical) | 57 | 99 | 0 | 0 | no | 31 |
| 10 | m | r | MCA (subcortical) | 49 | 91 | 1 | 1 | no | 12 |
| 11 | m | l | MCA (subcortical) | 57 | 99 | 1 | 0 | yes | 37 |
| 12 | m | r | Brainstem | 57 | 99 | 0 | 1 | no | 44 |
| 13 | f | r | MCA (cortical) | 56 | 91 | 3 | 0 | yes | 55 |
| 14 | m | r | MCA (cortical) | 57 | 99 | 4 | 0 | yes | 32 |
| 15 | m | l | MCA (subcortical) | 57 | 99 | 1 | 0 | yes | 43 |
| 16 | f | r | MCA (subcortical) | 57 | 76 | 2 | 1 | yes | 15 |
| 17 | m | r | Brainstem | 37 | 84 | 3 | 1 | yes | 82 |
| 18 | m | l | Brainstem | 44 | 83 | 1 | 1 | no | 30 |
| 19 | f | l | Brainstem | 56 | 76 | 1 | 1 | no | 12 |
| 20 | m | l | PCA (subcortical) | 55 | 92 | 1 | 0 | yes | 15 |
| 21 | m | l | MCA (subcortical) | 57 | 99 | 4 | 1 | yes | 33 |
| 22 | m | l | MCA (sub- & cortical) | 57 | 99 | 1 | 0 | yes | 25 |
| 23 | m | r | MCA (subcortical) | 53 | 99 | 1 | 0 | yes | 35 |
| 24 | f | r | Brainstem | 57 | 83 | 1 | 1 | no | 20 |
| 25 | m | r | MCA (subcortical) | 57 | 99 | 4 | 0 | yes | 23 |

**Supplementary Table 2: Correlation analyses between different aspects of motor control and cortico-cortical connections after tract-specific subject exclusion.** For three patients, distinct cortico-cortical tracts were excluded because lesions affected a considerable proportion (>10%) of the tract's one-directional voxels. Analyses were carried out separately for (i) basal and (ii) complex motor control. Partial correlations assessed the relationship between motor control and tractwise anisotropy while controlling for ipsilesional CST integrity. Bold font indicates significance after FDR-correction ($p < .05$). Asterisks signify the following significance thresholds: *** $p < .001$, ** $p < .01$, * $p < .05$.

| | **basal motor control** | | | |
|---|---|---|---|---|
| | **Pearson correlations** | | **partial correlations** | |
| **connection** | **r** | **p (FDR)** | **r** | **p (FDR)** |
| **homologous** | | | | |
| clM1-ilM1 | **0.62** | **0.002**** | **0.45** | **0.040*** |
| **intrahemispheric** | | | | |
| ilPMd-ilM1 | **0.66** | **0.002**** | **0.61** | **0.005**** |
| ilPMv-ilM1 | **0.74** | **<0.001***** | **0.67** | **0.003**** |
| ilSMA-ilM1 | **0.63** | **0.002**** | **0.65** | **0.003**** |
| **interhemispheric** | | | | |
| clPMd-ilM1 | 0.31 | 0.134 | 0.25 | 0.246 |
| clPMv-ilM1 | **0.56** | **0.007**** | **0.54** | **0.013*** |
| clSMA-ilM1 | **0.43** | **0.036*** | 0.31 | 0.172 |
| | **complex motor control** | | | |
| **homologous** | | | | |
| clM1-ilM1 | **0.49** | **0.022*** | 0.26 | 0.246 |
| **intrahemispheric** | | | | |
| ilPMd-ilM1 | **0.53** | **0.018*** | 0.45 | 0.073 |
| ilPMv-ilM1 | **0.60** | **0.015*** | 0.49 | 0.073 |
| ilSMA-ilM1 | **0.56** | **0.015*** | **0.56** | **0.035*** |
| **interhemispheric** | | | | |
| clPMd-ilM1 | 0.28 | 0.183 | 0.21 | 0.332 |
| clPMv-ilM1 | **0.46** | **0.029*** | 0.43 | 0.073 |
| clSMA-ilM1 | **0.45** | **0.029*** | 0.33 | 0.160 |

**Supplementary Table 3: Recovery-dependent subgroup analysis: Correlation analyses between basal motor control and cortico-cortical connections after tract-specific subject exclusion.** For three patients, distinct cortico-cortical tracts were excluded because lesions affected a considerable proportion (>10%) of the tract's one-directional voxels. Analyses were carried out separately for patients featuring (i) substantial or (ii) no substantial recovery as assessed by the difference in NIHSS-arm score in the acute and chronic stage. Partial correlations assessed the relationship between basal motor control and tractwise anisotropy while controlling for ipsilesional CST integrity. Bold font indicates significance after FDR-correction ($p < .05$). Asterisks signify the following significance thresholds: *** $p < .001$, ** $p < .01$, * $p < .05$.

| | **substantial recovery** | | | |
|---|---|---|---|---|
| | **Pearson correlations** | | **partial correlations** | |
| **connection** | **r** | **p (FDR)** | **r** | **p (FDR)** |
| **homologous** | | | | |
| cIM1-iIM1 | **0.79** | **0.003**** | **0.75** | **0.016*** |
| **intrahemispheric** | | | | |
| iIPMd-iIM1 | 0.31 | 0.397 | 0.33 | 0.387 |
| iIPMv-iIM1 | 0.62 | 0.079 | 0.58 | 0.166 |
| iISMA-iIM1 | 0.16 | 0.582 | 0.26 | 0.467 |
| **interhemispheric** | | | | |
| cIPMd-iIM1 | 0.22 | 0.492 | 0.18 | 0.549 |
| cIPMv-iIM1 | 0.35 | 0.390 | 0.36 | 0.387 |
| cISMA-iIM1 | 0.50 | 0.139 | 0.42 | 0.325 |
| | **no substantial recovery** | | | |
| **homologous** | | | | |
| cIM1-iIM1 | 0.44 | 0.279 | 0.17 | 0.657 |
| **intrahemispheric** | | | | |
| iIPMd-iIM1 | **0.85** | **0.007**** | 0.77 | 0.053 |
| iIPMv-iIM1 | **0.81** | **0.010*** | 0.73 | 0.060 |
| iISMA-iIM1 | **0.90** | **0.003**** | **0.87** | **0.015*** |
| **interhemispheric** | | | | |
| cIPMd-iIM1 | 0.33 | 0.392 | 0.34 | 0.521 |
| cIPMv-iIM1 | **0.71** | **0.036*** | 0.68 | 0.076 |
| cISMA-iIM1 | 0.30 | 0.392 | 0.21 | 0.657 |
